# A spatial model to optimise predictions of COVID-19 incidence risk in Belgium using symptoms as reported in a large-scale online survey

**DOI:** 10.1101/2020.05.18.20105627

**Authors:** Thomas Neyens, Christel Faes, Maren Vranckx, Koen Pepermans, Niel Hens, Pierre Van Damme, Geert Molenberghs, Jan Aerts, Philippe Beutels

## Abstract

Although COVID-19 has been spreading throughout Belgium since February, 2020, its spatial dynamics in Belgium remain poorly understood, due to the limited testing of suspected cases. We analyse data of COVID-19 symptoms, as self-reported in a weekly online survey, which is open to all Belgian citizens. We predict symptoms’ incidence using binomial models for spatially discrete data, and we introduce these as a covariate in the spatial analysis of COVID-19 incidence, as reported by the Belgian government during the days following a survey round. The symptoms’ incidence predictions explain a significant proportion of the variation in the relative risks based on the confirmed cases, and exceedance probability maps of the symptoms’ incidence and the confirmed cases’ relative risks pinpoint the same high-risk region. We conclude that these results can be used to develop public monitoring tools in scenarios with limited lab testing capacity, and to supplement test-based information otherwise.

## 1. Introduction

COVID-19 is a respiratory disease caused by a highly infectious single-stranded RNA corona virus, SARS-CoV-2 (Chen *et al*. 2020, Wu *et al*. 2020). It was first observed in Wuhan, the capital of the Hubei province in the People’s Republic of China, in December 2019 (Zhu *et al*. 2020). The virus most likely has a zoonotic origin, but human-to-human transmission, which happens mainly via droplets and fomites, combined with a high basic reproductive number, has caused the disease to rapidly spread across continents. It has been declared a global pandemic on March 11, 2020 (WHO 2020).

The first imported COVID-19 case in Belgium was reported on February 4, 2020, in Brussels; this case did not lead to further infections. Due to various further introductions, the disease spread throughout the country. The Belgian government has undertaken several measures to slow down community transmission, the most notable of which has been the implementation of a lockdown of the country on March 18, 2020. Due to limited capacity, only a fraction of suspected Belgian COVID-19 patients has been tested to confirm SARS-CoV-2 infection. These are primarily severe cases, which has complicated the assessment of the true extent of the disease’s spatio-temporal spread.

The University of Antwerp, in collaboration with Hasselt University and KU Leuven, has designed an ethically approved weekly online COVID-19 survey (https://www.uantwerpen.be/en/projects/corona-study/), which is open to the general Belgian public. A key objective of the survey is to collect information on COVID-19 symptoms from the general public. The weekly number of participants has been large; during its first four rounds, the survey reached 537,172; 334, 935; 397, 529; and 215,138 respondents, respectively, with complete residential and personal information to conduct a spatial analysis. However, as the survey may not reach all segments of society equally (Alessi & Martin 2010; Andrews *et al*. 2003), it remains unclear whether sampling bias invalidates statistical inference on the spatial dynamics of COVID-19-like symptoms as a proxy for the distribution of COVID-19.

Geostatistical models are often applied to analyse and predict disease risk in a population (Diggle & Robeiro 2007). Using methods for spatially discrete outcomes (Besag *et al*. 1991; Lawson 2013), we can predict COVID-19 incidence via crowd-sourced data of symptoms obtained by self-reporting in the online survey. We can use these predictions to optimally model the geographical risk distribution of confirmed COVID-19 cases, as reported by the Belgian government. This additionally allows to investigate routes to develop an early-warning framework aimed at detecting COVID-19 cases by self-reporting citizens, when large-scale testing and tracing of the general public is not feasible, and to supplement information obtained from testing and tracing otherwise.

In this study, we fit spatial models to data obtained during the third round of the online survey, conducted on March 31, 2020, and data of confirmed cases, as reported by the Belgian population health institute (Sciensano) between April 7 and April 9, 2020. We use approximate Bayesian estimation methods to spatially analyse self-reported COVID-19 symptoms. We then use mean incidence predictions as a plug-in covariate in a spatial model to analyse confirmed cases. Our aim is to investigate whether symptoms that are reported in an online survey, which is ordinarily subject to sampling bias, are useful to predict the spatial spread of detected COVID-19 disease approximately one week later.

## 2. Methodology

### 2.1. Data

We make use of Belgian data of 5183 confirmed COVID-19 cases with known residential, age, and gender information, as reported by the Belgian population health institute on April 7, April 8, and April 9, 2020 (henceforth, *covid data*). Fig. 1 depicts the standardized incidence rates, *SIR_i_* = *O_i_/E_i_*, with *O_i_* and *E_i_* the observed number of cases and the internally age-gender standardised expected counts, respectively, for municipality *i* = 1,…, 589, using data of all confirmed cases between April 7 and April 9, 2020, in Belgium. We use age groups in the standardisation process, more specifically the age intervals, 0 − 24, 25 − 44, 45 − 64, and +65 years old. The widths of the age intervals are based on considerations related to the online survey data set; more information is provided in Section 2.2. Note that on Jan 1, 2019, a number of Belgian municipalities have been geographically and administratively united, which reduced the total number of Belgian municipalities from 589 to 581. We use the Belgian municipality structure of 2018 to improve spatial resolution, along with demographical information from the same year, which differs only minimally from the demography in 2020.

**Figure 1:**
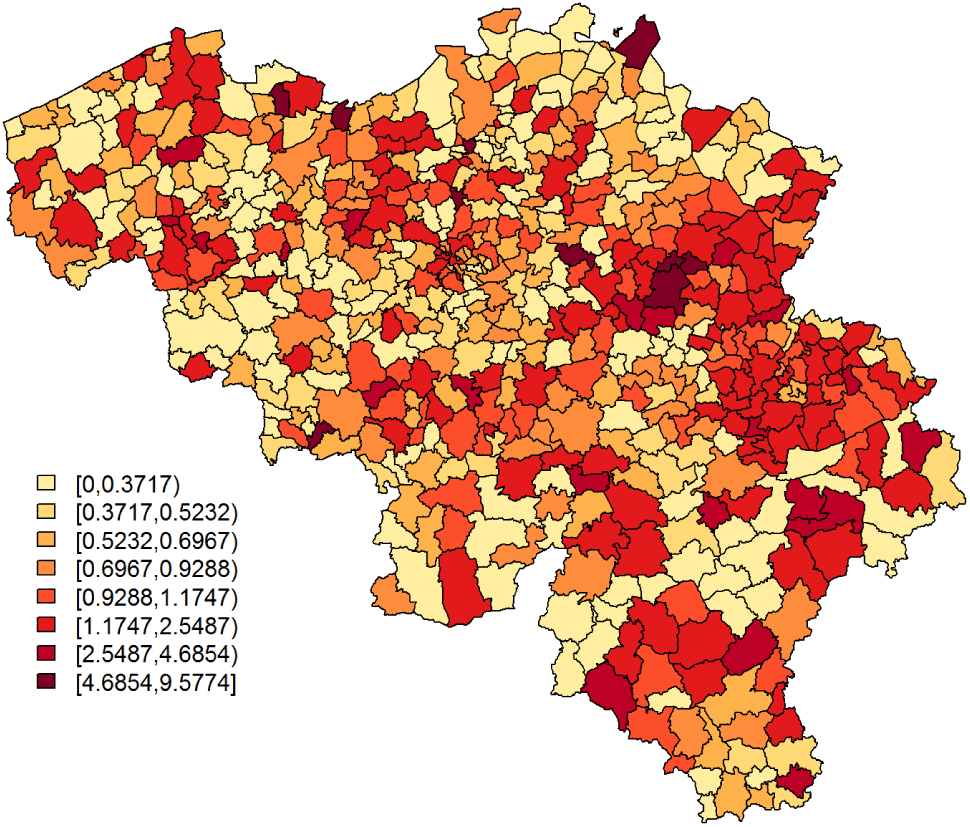
SIR of COVID-19 cases per municipality, based on all confirmed cases between April 7 and April 9, 2020.

Secondly, we use data on COVID-19 symptoms, as self-reported by participants in the third round of the online COVID-19 survey (March 31, 2020; henceforth, *symptoms data*), for which all necessary ethical approvals have been obtained. The survey can be filled in by all members of the public and is designed to collect data about spatial trends in COVID-19 symptoms within Belgium, the extent to which members of the public adhere to measures taken by the government, contact behaviour, and mental health dynamics, among others. We investigate data of the third round in the main analysis presented here, since (i) the survey in round 1 only contained one general question that gauged whether individuals experienced any flu-like symptoms. From round 2 onwards, this question was replaced by thirteen separate questions regarding specific COVID-19 symptoms; (ii) of the remaining surveys, round 3 had the largest sample size and the best coverage in Wallonia, the southern part of Belgium; (iii) during rounds 1 and 2, there was considerable overlap with the end of the influenza season, while exploratory analyses of symptom shifts through time signal the start of the pollen allergy season in round 4. We provide analysis results of survey rounds 2 and 4 as an Appendix (Section 6.2). We use data of males and females - not intersex due to the category’s limited sample size - with available age and residential information. Note that the online survey collected residential information on the postal code level, a subdivision of the municipality level. This yields 397, 529 data records for 1083 out of the 1133 Belgian postal code areas, with at least one respondent from each of the 589 Belgian municipalities. The majority of the respondents comes from Flanders, the northern part of Belgium (Fig. 2). All participants were asked to indicate which of the following COVID-19-like symptoms they experienced during the week preceding the online survey (March 24 – 30, 2020), if any: (i) a rapidly increasing fever, (ii) a high fever, (iii) a dry cough, (iv) shortness of breath, (v) chest pain, (vi) muscle pain, (vii) exhaustion, (viii) chills, (ix) nausea, (x) painful eyes, (xi) a sore throat, (xii) a rattling cough, and/or (xiii) a running nose. A binary variable *Y_jk_* takes a value 1 when person *k* = 1,…, *n_j_* in postal code *j* = 1,…, 1133 experienced at least one of the most typical symptoms, which we define as symptoms (i)-(iv), based on Jiang *et al*. (2020), Yang *et al*. (2020), and WHO (2020); otherwise, *Y_jk_* = 0.

**Figure 2:**
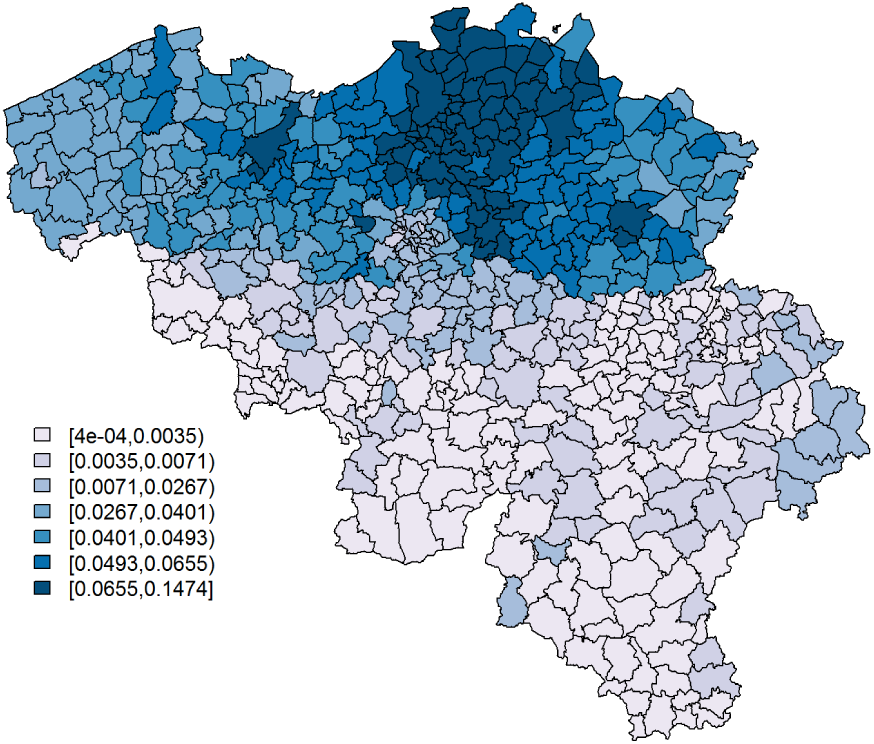
The proportion of the population per municipality taking the survey on March 31, 2020.

### 2.2. Statistical methods

We fit conditional autoregressive (CAR) convolution models (Besag *et al*. 1991) to the *symptoms* and *covid* data, using integrated nested Laplace approximation (INLA, Rue *et al*. 2009). INLA is a convenient approximate Bayesian estimation method that computes approximations of posterior marginal distributions for latent Gaussian models. We apply it in R 4.0.0 (R Core Team 2020), through the package R-INLA.

For the *symptoms* data, the CAR convolution model is given by

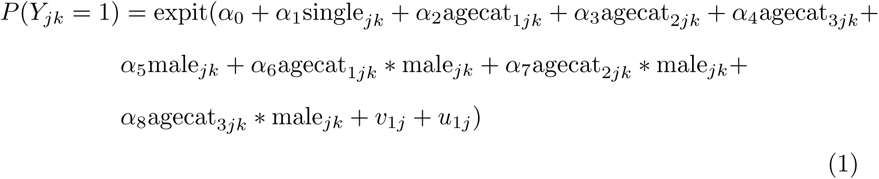

where *single* denotes a binary variable taking the value 1 when a participant is the only member of a household and 0 otherwise; *agecat* _1_, *agecat*_2_, and *agecat*_3_ are dummy variables that indicate whether participants belong to the age groups 25 − 44, 45 − 64, and +65, respectively, the interval widths of which we have chosen to categorise the data into groups expected to showcase different social behaviour, while maintaining balanced sample sizes among these categories; *male* = 1 for males, 0 for females. We correct for spatially uncorrelated heterogeneity on the postal code level (UH) with a normally distributed random effects term:

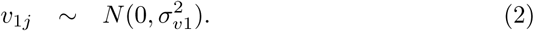

Spatially correlated heterogeneity (CH) is accommodated by *u_j_*, an intrinsic conditional autoregressive (CAR) random effects term, such as introduced by Besag & Kooperberg (1995),

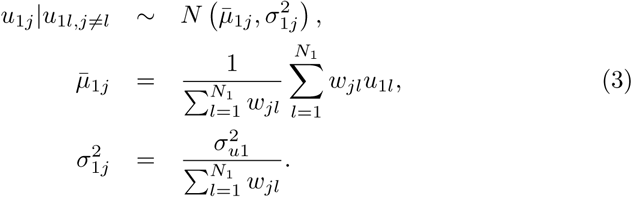

Here, *W_jl_* = 1 if areas *j* and *l* are adjacent and 0 otherwise, and N_1_ = 1133. We then predict 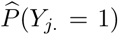, the predicted probability of a postal code area’s inhabitant to experience at least 1 typical COVID-19 symptom, which is corrected for the age, gender, and single households dynamics of the municipality in which the postal code area is situated, since this demographic information on the postal code level is not at our disposal.

For the *covid* data, we fit a Poisson CAR convolution model,

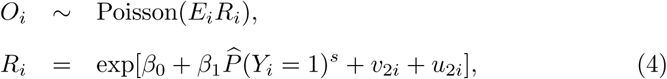

where *R_i_* denotes the relative risk for municipality *i*. *v*_2_*_i_* and *u*_2_*_i_* are defined similarly as *v*_1_*_j_* and *u*_1_*_j_* in (2) and (3), respectively, but with different separate heterogeneity terms denoted by 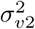, 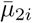,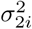, 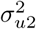, and *N*_2_ = 589 instead of 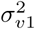, 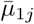, 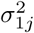, 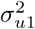, and *N*_1_, respectively. The probability of a municipality’s inhabitant to experience at least 1 typical COVID-19 symptom is calculated as,

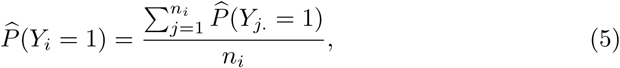

with *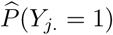* as predicted by (1), and where *n_i_* denotes the number of postal code areas that municipality *i* consists of. We include it in its standardised form, which we denote as 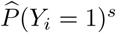, as a risk factor in the model.

We use vague priors: *N*(0,1000) for all covariate effects, and Gamma(1,0.0005), here parameterised with a shape and rate parameter, for 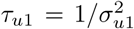, 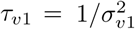, 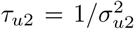, and 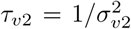. A sensitivity analysis for the choice of the prior distribution, where we use Gamma(1,0.01) as a prior for the precision parameters, has been documented in the Appendix (Section 6.1). Although the use of these priors affects the precision estimates, the estimates of covariate effects and the maps displaying predictions and exceedance probabilities, remain almost unchanged. We denote covariate effects as *significant*, when their associated 95% credible interval does not include 0.

## 3. Results

The upper panel of Table 1 reports parameter estimates for the analysis of the *symptoms* data. Being single is significantly associated with a lower probability to report at least 1 typical COVID-19 symptom. However, its effect is small. Age and gender have significant interaction effects; we see the largest probability among non-single females between 25 and 44 years old, while the lowest probability is seen in single elderly males. Note that the UH random effects explain little variability. We do not remove it from the model, since we would then assume that there is no small-scale extra-variability. Fig. 3 and 4 show, respectively, 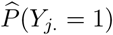, after correcting for demographic variation in age, gender, and household, i.e., singles vs. non-singles, and the exceedance probabilities, 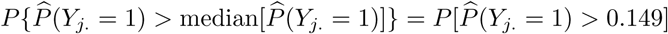.

**Table 1:**
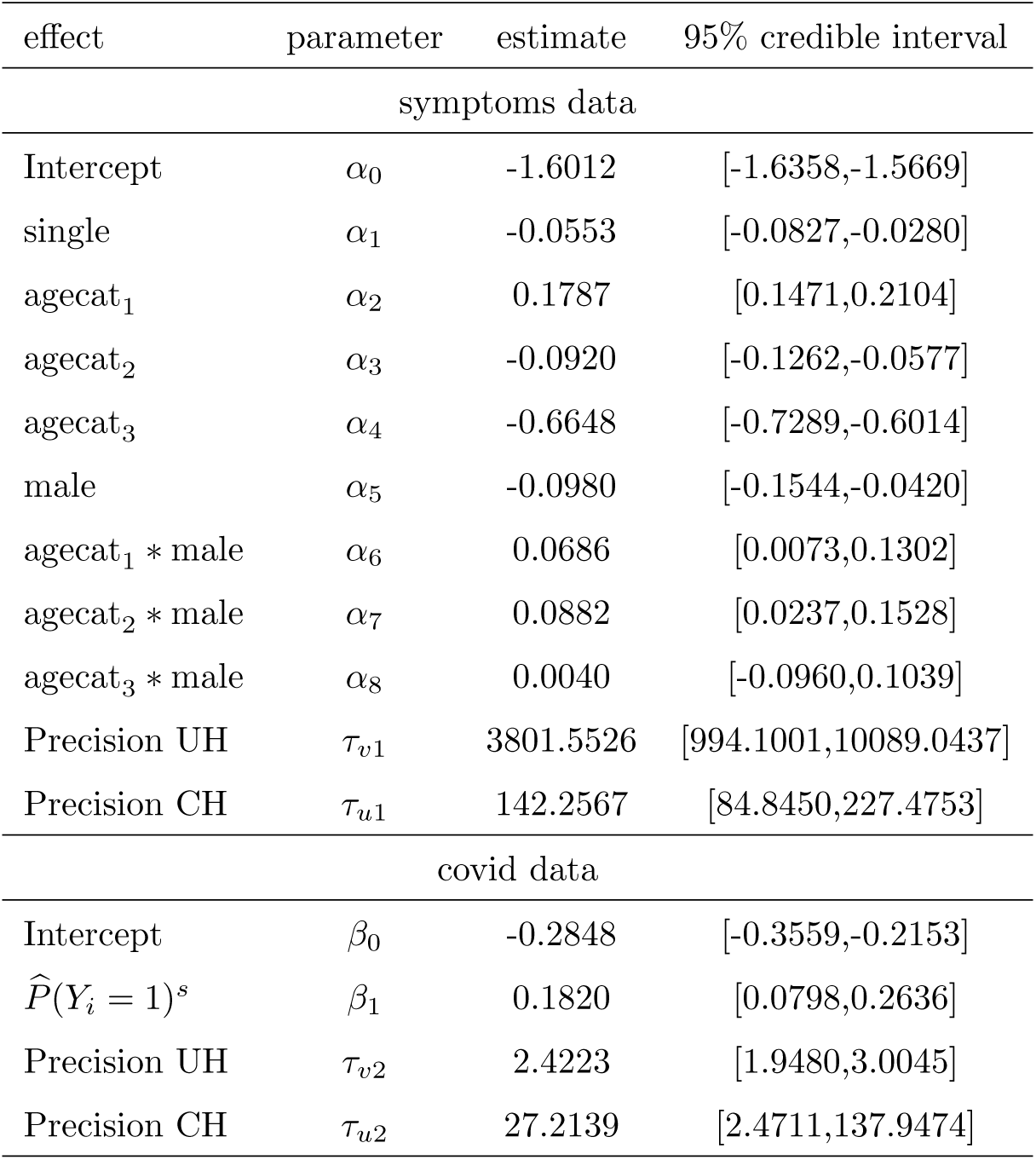
Estimation results.

**Figure 3:**
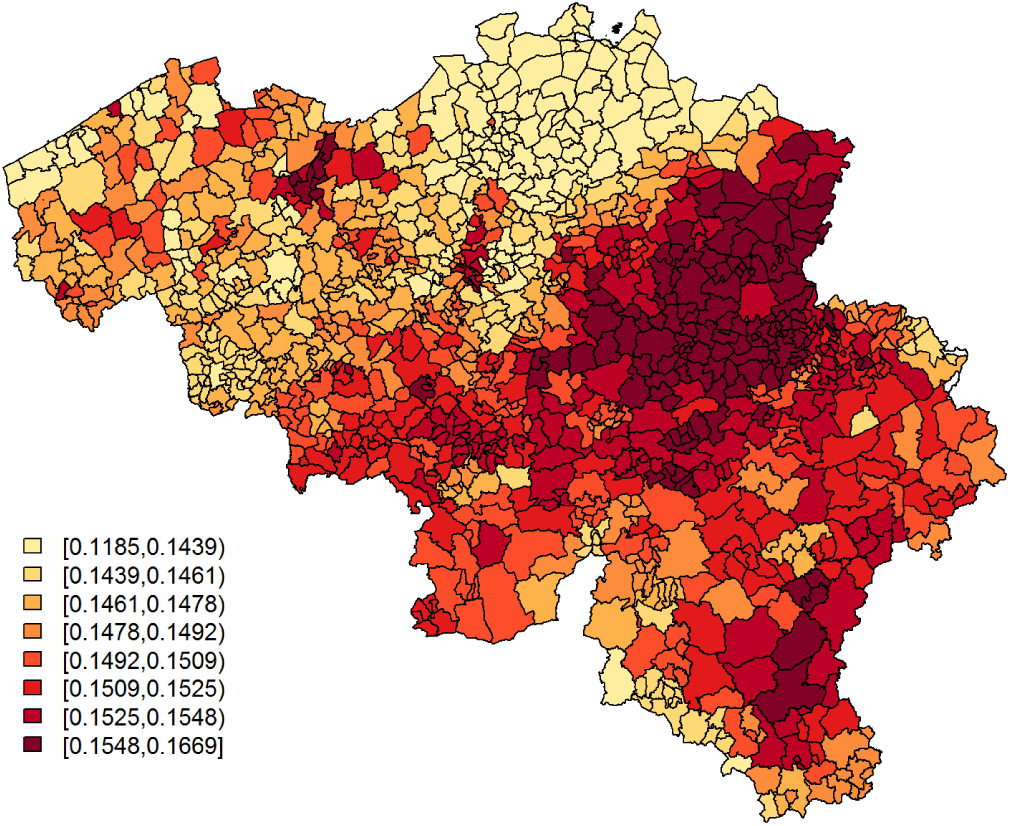
Predicted probabilities for a citizen to experience at least 1 of 4 typical COVID-19 symptoms per postal code area.

**Figure 4:**
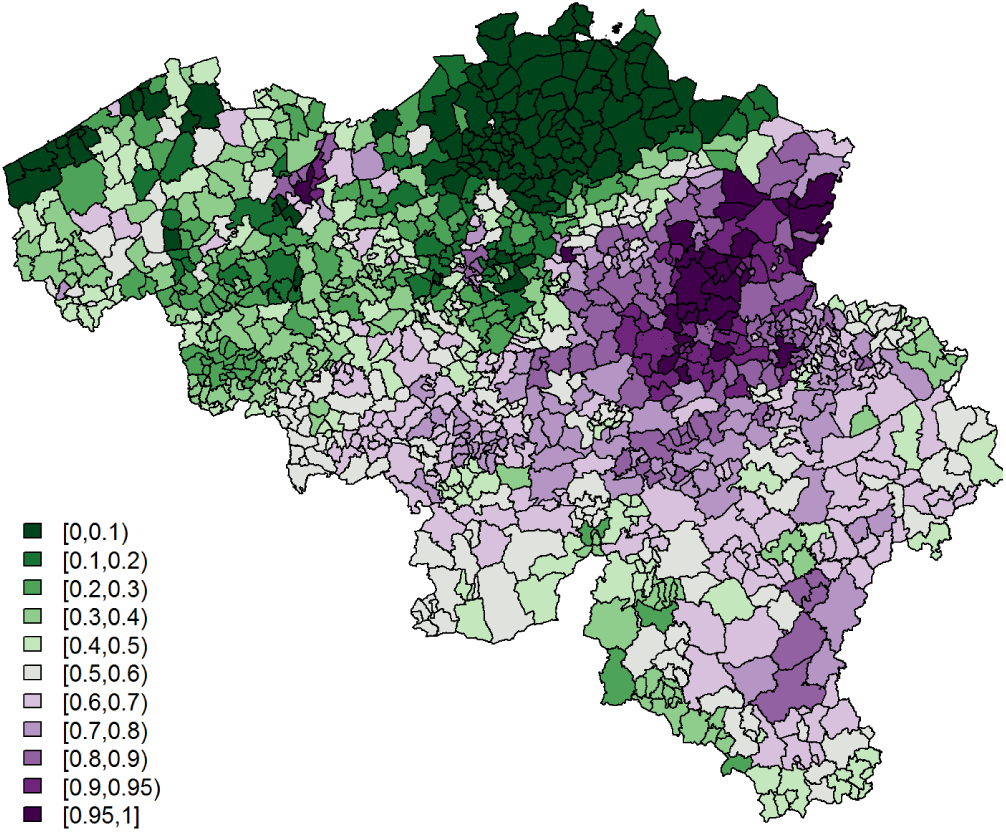
Exceedance probabilities per postal code area for the predicted probability for a citizen to experience at least 1 of 4 typical COVID-19 symptoms, with threshold = 0.149.

The lower panel of Table 1 presents parameter estimates for the *covid* data analysis. The symptoms’ incidence, as predicted by (1), is significantly and positively associated with the relative incidence risk, based on the confirmed cases. Fig. 5 and 6 show, respectively, 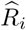 and the exceedance probabilities, 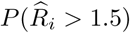. The cluster of postal code areas with elevated predicted incidence of typical COVID-19 symptoms in the central-east of Belgium, situated around the city of Sint-Truiden, in Fig. 4, overlaps well with the cluster of municipalities in Fig. 6 that has a high probability, i.e., 95%, to have at least a 150% increase in relative incidence risk.

**Figure 5:**
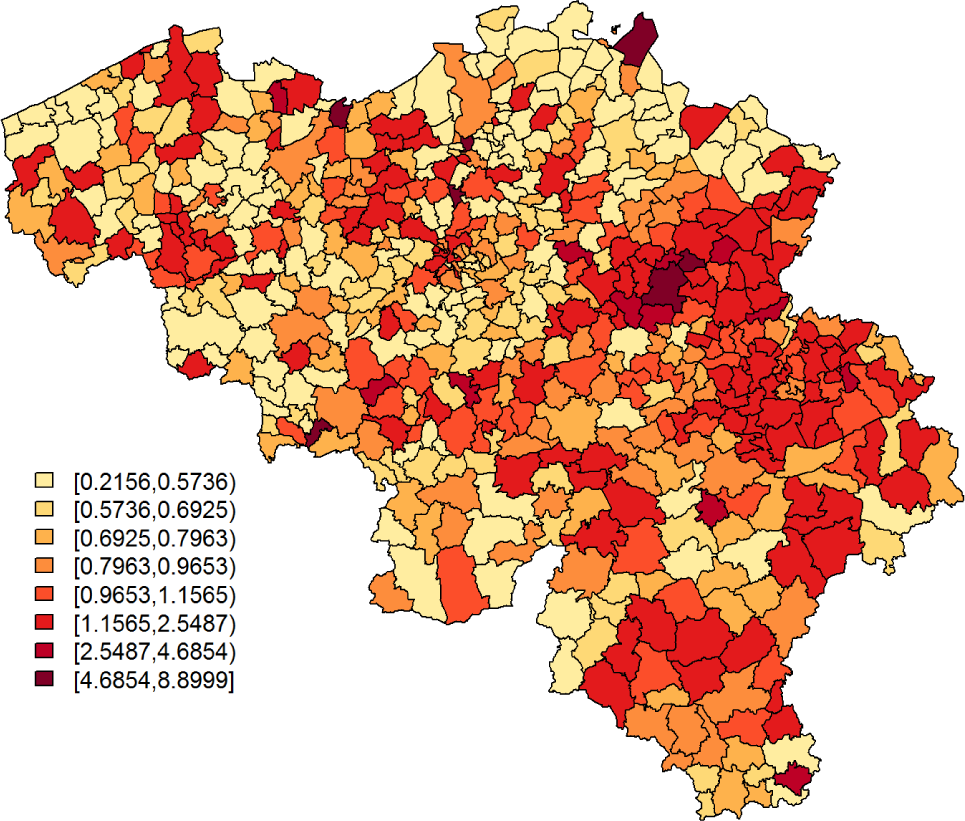
Predicted COVID-19 relative risk per municipality, based on data of confirmed cases between April 7 and April 9, 2020.

**Figure 6:**
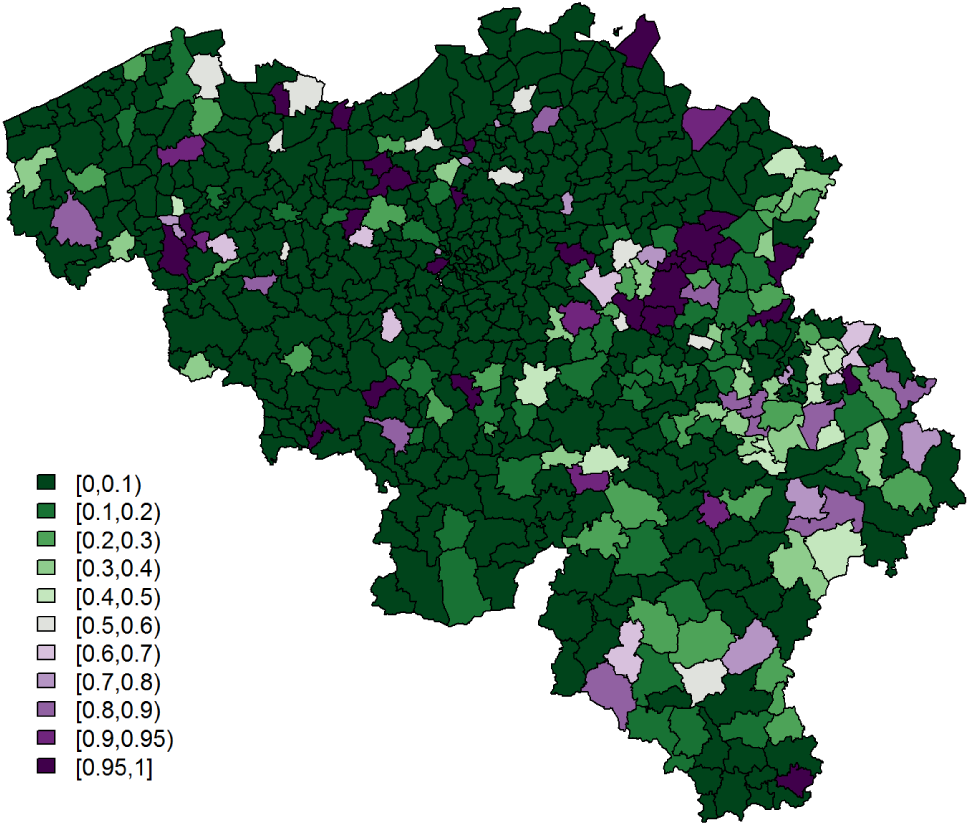
Exceedance probabilities for the relative risk per municipality, based on data of confirmed cases between April 7 and April 9, 2020, with relative risk threshold = 1.5.

The symptoms that were self-reported to be experienced during the period of March 24 − 30, have a significant predictive effect on the incidence risk of confirmed cases within a period that spans more days than the period of April 7 – 9 (Table 2). We find significant results for the effect of 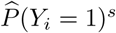 on the incidence risk of confirmed cases for all three-day periods throughout April 2 and April 10, and a borderline significant effect when analysing all cases that were confirmed between April 2 and April 10 together. Based on the effect size and the credible intervals’ widths, the optimal predictive performance is suggested for the period between April 7 and April 9. Although it is uncommon to consider traditional issues related to multiple testing in the context of a Bayesian analysis, we note that for results that we consider as borderline significant, signals might reflect spurious correlations.

**Table 2:**
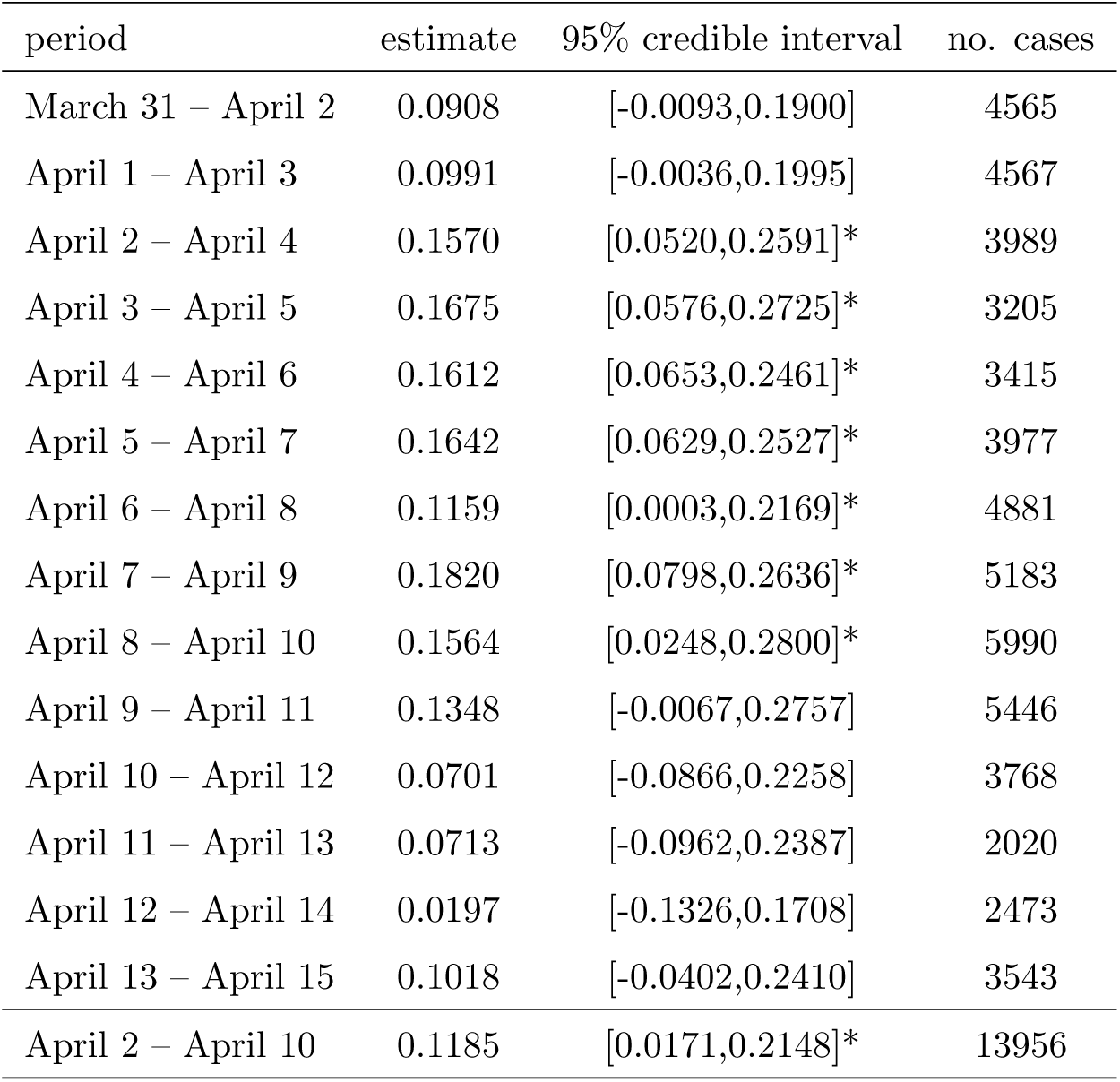
Estimation results for *β*_1_, the effect of 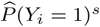, when investigating different time periods of confirmed cases. An asterisk (*) denotes a significant effect on a 5% significance level.

## 4. Discussion and conclusion

Our study shows that, when using geographical crowd-sourced information on COVID-19 that is obtained by self-reporting within a large-scale survey study, model-based symptom incidence predictions are capable of explaining a significant proportion of the heterogeneity that is seen in the number of confirmed COVID-19 cases, as reported by the government, within 3 to 18 days after these symptoms were experienced. Moreover, exceedance probabilities, based on the analysis of the *symptoms* data, pinpoint an important cluster of elevated COVID-19 risk around the city of Sint-Truiden in central eastern Belgium, which aligns well with a region that has since then received increased attention, due to a number of local outbreaks.

Note that we have conducted the same analyses, using *symptoms* data from rounds 2 and 4 of the online survey, which we document as an appendix (Section 6.2). Similarly as in the analysis based on survey 3, the predictive means of the symptoms’ incidence significantly explain variation in the number of confirmed cases, but for a more restricted set of three-day periods within a 14-day time span after the day of the respective surveys. Their predictive performances are weaker than those obtained from survey 3 and it is less clear to pinpoint a time frame for which the survey data optimally predict confirmed cases. As explained in Section 2.1, this is likely due to a combination of the overlapping influenza and allergy season during rounds 2 and 4, respectively, and a lower amount of participants from Wallonia, which may obstruct the detection of all spatial dynamics of COVID-19 symptoms in Belgium. Note that the *symptoms* data reflect symptoms that were experienced during the week preceding the respective rounds of the survey. This period does not necessarily reflect the moment of the symptoms’ onsets, which may have taken place earlier. Future studies will investigate how data of COVID-19 symptoms’ onsets can be optimally linked to data of confirmed cases.

Note that, for the data from round 3 of the survey, not all high-risk areas in Fig. 6 could be detected by the *symptoms* data analysis (Fig. 4). We believe that this might be due to a considerably large amount of small-scale variation in virus transmission, such that our model is currently best at pinpointing clusters that are occurring on a moderately large scale. This might be explained by the quarantine measures that obstruct typical transmission routes, such that infection mostly occurs very localised (Ganyani *et al*. 2020). However, what is defined as small-scale spatial variation depends on the spatial resolution of data; in order to develop COVID-19 monitoring tools, analysts will need spatial information on finer scales than the municipality or postal code level. Furthermore, similarly to the previous conclusions with respect to rounds 2 and 4 of the survey, the relatively small sample sizes in the southern part of Belgium for the *symptoms* data likely hamper the detection of high-incidence areas in the analysis of round 3 as well. This highlights the need for investments in monitoring tools, and promotional campaigns to engage the general public throughout the whole region to participate in these online surveys, in combination with such other measures as, for example, tracing strategies.

With respect to demographic heterogeneity in typical COVID-19 symptoms, the analysis results report the lowest symptoms’ incidence for elderly, while symptoms occur mostly among persons between 25 and 44 years old, especially females with at least one additional household member. These differences might be a result of variation in the social distancing behaviour between age groups. Preliminary analyses of contact behaviour, based on the online survey data (not shown), suggest that Belgian elderly started to engage in social distancing significantly sooner than younger individuals during the COVID-19 outbreak; among individuals who are younger than 65 years, those younger than 25 are suggested to be the slowest to adapt to social distancing measures. A plausible reason why the latter is not reflected in the *symptoms* data analysis results, is that among COVID-19 patients, children and adolescents in general are less likely to experience typical COVID-19 symptoms (Dong *et al*. 2020).

This study can be improved by investigating the outcomes spatio-temporally. However, the correct extraction of the specific day of the symptoms’ onset from the online survey data should be undertaken with care and will be investigated in the future. We have therefore analysed symptoms data that were aggregated in time. Moreover, we plan to develop a joint modelling framework in which we simultaneously model symptoms and confirmed cases, e.g., by extending correlated random-effects models proposed by Neyens *et al*. (2016). This will allow us to exploit spatial dependence that is likely to occur between symptoms’ incidence and the confirmed cases’ disease risk. This can improve the current two-step approach where we use model-based symptom predictions as a plug-in covariate to model the spatial dynamics of the confirmed cases’ disease risk.

## Data Availability

The data of confirmed COVID-19 cases, as analysed in this study, are confidential. They are available in an aggregated format on https://epistat.wiv-isp.be/covid/. The data of COVID-19 symptoms that were reported by participants of the Big Corona Study are confidential, but can be accessed after approval by the Big Corona Study steering committee.

## Acknowledgements

We thank Herman Van Oyen and Toon Braeye from the Belgian population health institute (Sciensano) for reading and commenting on our manuscript.

## Funding

This research received funding from the Flemish Government (AI Research Program). Authors NH and PB acknowledge funding from the European Union’s Horizon 2020 research and innovation programme - project EpiPose (No 101003688).

## Declaration of interest

The authors declare no conflicts of interest.

## Data availability statement

The *covid* data, as analysed in this study, are confidential. They are available in an aggregated format on https://epistat.wiv-isp.be/covid/. The *symptoms* data are confidential, but can be accessed after approval by the Corona Study steering committee.

## Ethics statement

All data were collected in ethically approved studies.

## Author contributions

Thomas Neyens: Conceptualization, Methodology, Software, Writing-Original draft preparation. Christel Faes: Methodology, Writing-Reviewing and Editing. Maren Vranckx: Formal Analysis, Software. Koen Pepermans: Investigation, Resources, Writing-Reviewing and Editing. Niel Hens: Supervision, Writing-Reviewing and Editing. Pierre Van Damme: Resources, Writing-Reviewing and Editing. Geert Molenberghs: Conceptualization, Writing-Reviewing and Editing. Jan Aerts: Data curation, Writing-Reviewing and Editing. Philippe Beutels: Investigation, Resources, Supervision, Writing-Reviewing and Editing.

## 6. Appendix

### 6.1. Sensitivity analysis

In the sensitivity analysis, we use Gamma(1,0.01) instead of Gamma(1,0.0005) as a prior for the precision parameters for the UH and CH random effects terms.

**Table 3:** Estimation results.

| effect | parameter | estimate | 95% credible interval |
| --- | --- | --- | --- |
| symptoms data |  |  |  |
| Intercept | $\alpha_0$ | -1.6018 | [-1.6364,-1.5673] |
| single | $\alpha_1$ | -0.0556 | [-0.0831,-0.0283] |
| agecat<sub>1</sub> | $\alpha_2$ | 0.1786 | [0.1470,0.2103] |
| agecat<sub>2</sub> | $\alpha_3$ | -0.0920 | [-0.1262,-0.0577] |
| agecat<sub>3</sub> | $\alpha_4$ | -0.6649 | [-0.7289,-0.60142] |
| male | $\alpha_5$ | -0.0980 | [-0.1544,-0.0420] |
| agecat<sub>1</sub> * male | $\alpha_6$ | 0.0686 | [0.0072,0.1302] |
| agecat<sub>2</sub> * male | $\alpha_7$ | 0.0881 | [0.0237,0.1528] |
| agecat<sub>3</sub> * male | $\alpha_8$ | 0.0041 | [-0.0960,0.1040] |
| Precision UH | $\tau_{v1}$ | 666.2017 | [375.0517,1099.2037] |
| Precision CH | $\tau_{u1}$ | 148.0921 | [88.0472,235.2848] |
| covid data |  |  |  |
| Intercept | $\beta_0$ | -0.2849 | [-0.3559,-0.2155] |
| $\hat{P}(Y_i = 1)^s$ | $\beta_1$ | 0.1804 | [0.0781,0.2621] |
| Precision UH | $\tau_{v2}$ | 2.4286 | [1.9496,3.0140] |
| Precision CH | $\tau_{u2}$ | 25.0809 | [2.4016,123.9483] |

**Figure 7:**
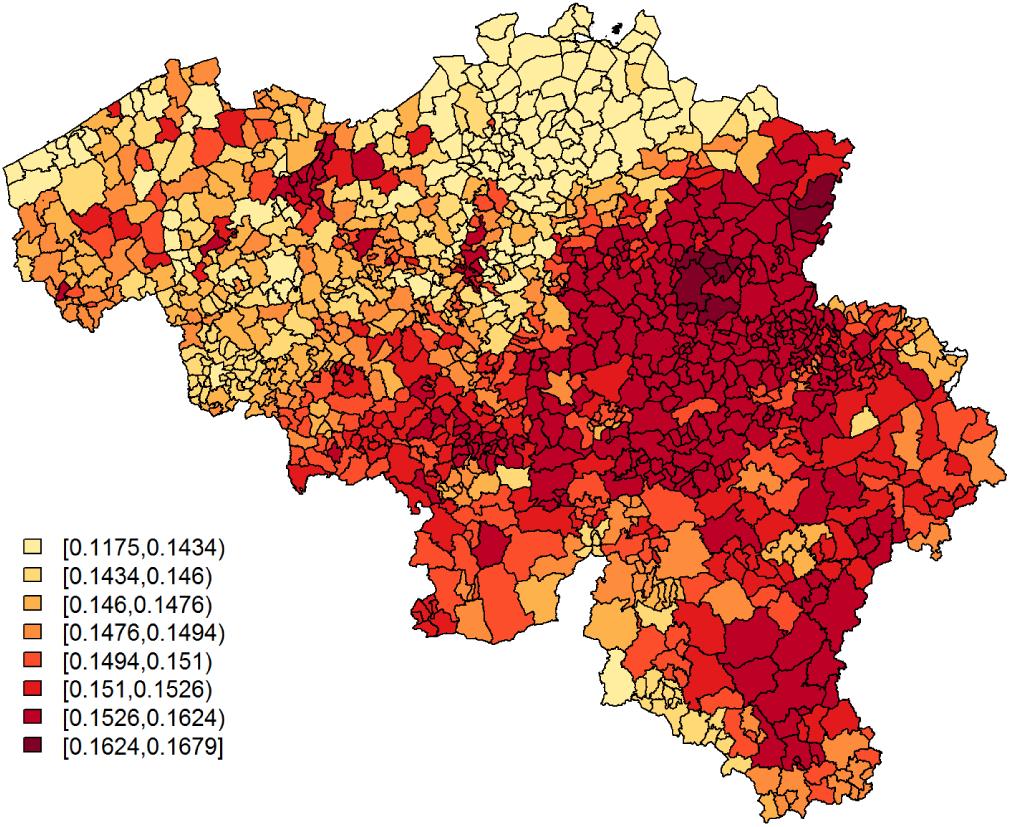
Sensitivity analysis - Predicted probabilities for a citizen to experience at least 1 of 4 typical COVID-19 symptoms per postal code area.

**Figure 8:**
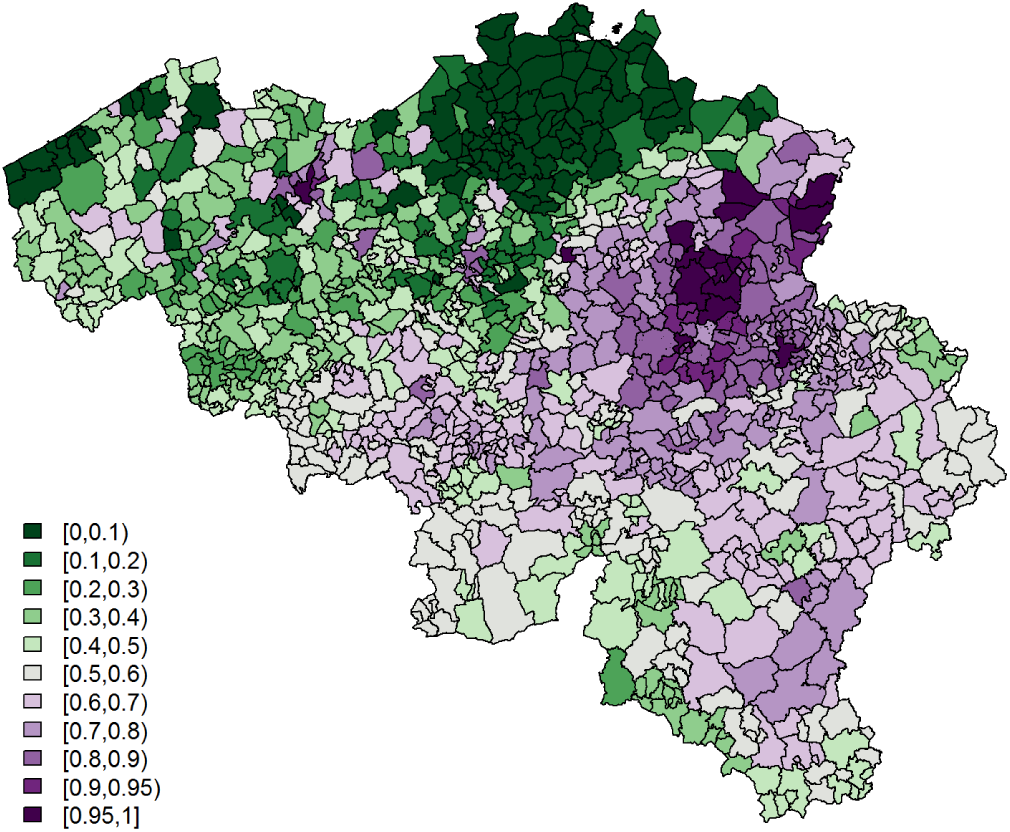
Sensitivity analysis - Exceedance probabilities per postal code area for the predicted probability for a citizen to experience at least 1 of 4 typical COVID-19 symptoms, with threshold = 0.149.

**Figure 9:**
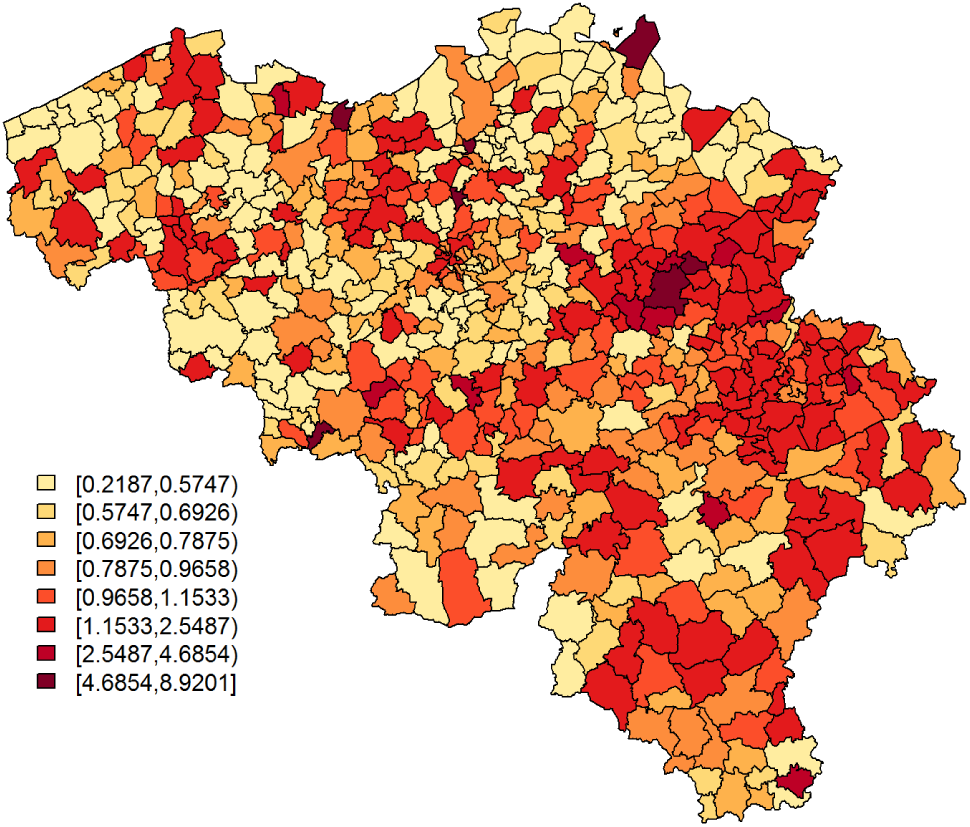
Sensitivity analysis - Predicted COVID-19 relative risk per municipality, based on data of confirmed cases between April 7 and April 9, 2020.

**Figure 10:**
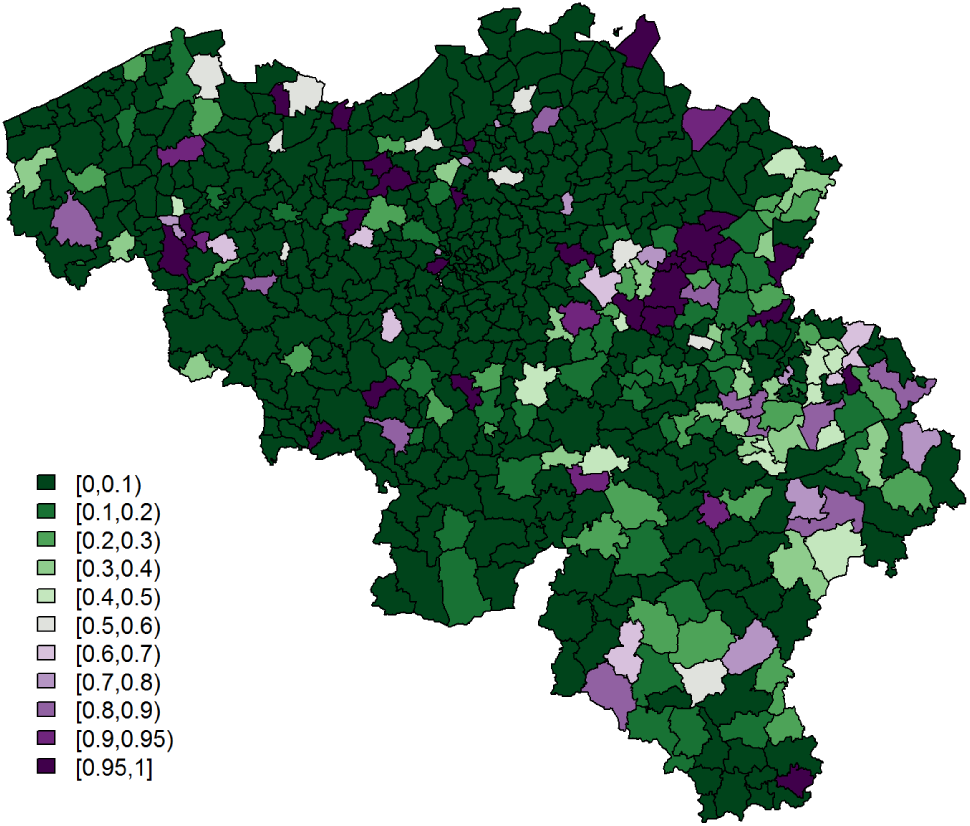
Sensitivity analysis - Exceedance probabilities for the relative risk per municipality, based on data of confirmed cases between April 7 and April 9, 2020, with relative risk threshold = 1.5.

### 6.2. Analyses of rounds 2 and 4

**Table 4:** Estimation results for *β*_1_, the effect of 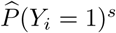, obtained from the analysis of 341, 320 respondents in the second round of the online survey, when investigating different time periods of confirmed cases. An asterisk (*) denotes a significant effect on a 5% significance level.

| period | estimate | 95% credible interval | no. cases |
| --- | --- | --- | --- |
| March 24 - March 26 | 0.1109 | [0.0211,0.2003]* | 3705 |
| March 25 - March 27 | 0.0543 | [-0.0439,0.1523] | 4016 |
| March 26 - March 28 | 0.0691 | [-0.0353,0.1732] | 3663 |
| March 27 - March 29 | 0.0610 | [-0.0473,0.1687] | 2987 |
| March 28 - March 30 | 0.0722 | [-0.0325,0.1756] | 3206 |
| March 29 - March 31 | 0.0947 | [-0.0102,0.1988] | 4039 |
| March 30 - April 1 | 0.0481 | [-0.0568,0.1523] | 4848 |
| March 31 - April 2 | 0.0801 | [-0.0264,0.1858] | 4565 |
| April 1 - April 3 | 0.0591 | [-0.0490,0.1653] | 4567 |
| April 2 - April 4 | 0.1068 | [-0.0053,0.2166] | 3989 |
| April 3 - April 5 | 0.1231 | [0.0047,0.2367]* | 3205 |
| April 4 - April 6 | 0.0903 | [-0.0196,0.1933] | 3415 |
| April 5 - April 7 | 0.1043 | [-0.0062,0.2086 ] | 3977 |
| April 6 - April 8 | 0.0609 | [-0.0593,0.1729] | 4881 |

**Table 5:** Estimation results for *β*_1_, the effect of 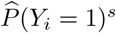, obtained from the analysis of 217877 respondents in the fourth round of the online survey, when investigating different time periods of confirmed cases. An asterisk (*) denotes a significant effect on a 5% significance level.

| period | estimate | 95% credible interval | no. cases |
| --- | --- | --- | --- |
| April 7 - April 9 | 0.2094 | [0.1303,0.2855]* | 5183 |
| April 8 - April 10 | 0.1986 | [0.0594,0.3272]* | 5990 |
| April 9 - April 11 | 0.1243 | [-0.0329,0.2813] | 5446 |
| April 10 - April 12 | 0.0753 | [-0.0959,0.2461] | 3768 |
| April 11 - April 13 | 0.1775 | [-0.0092,0.3629] | 2020 |
| April 12 - April 14 | 0.1259 | [-0.0407,0.2892] | 2473 |
| April 13 - April 15 | 0.1483 | [-0.0069,0.2986] | 3543 |
| April 14 - April 16 | 0.0998 | [-0.0614,0.2608] | 4643 |
| April 15 - April 17 | 0.1346 | [-0.0295,0.2940] | 4532 |
| April 16 - April 18 | 0.1602 | [-0.0147,0.3266] | 3652 |
| April 17 - April 19 | 0.2057 | [0.0253,0.3695]* | 2468 |
| April 18 - April 20 | 0.2325 | [0.0676,0.3875]* | 2345 |
| April 19 - April 21 | 0.1419 | [-0.0089,0.2880] | 2867 |
| April 20 - April 22 | 0.1364 | [-0.0105,0.2781] | 3179 |

## Notes

### Competing Interest Statement

The authors have declared no competing interest.

## References

Alessi, E. J., & Martin, J. I. (2010) Conducting an Internet-based Survey: Benefits, Pitfalls, and Lessons Learned. Social Work Research, 34, 122–128.

Andrews, D., Nonnecke, B., & Preece, J. (2003) Electronic survey methodology: A case study in reaching hard-to-involve Internet users. International Journal of Human-Computer Interaction, 16, 185–210.

Besag, J. & Kooperberg, C. (1995) On conditional and intrinsic autoregressions. Biometrika, 82, 733–746.

Besag, J., York, J., & Mollie, A. (1991) Bayesian image restoration with two applications in spatial statistics. Annals of the Institute of Statistical Mathematics,43, 1–59.

Chen, N., Zhou, M., Dong, X., Qu, J., Gong, F., Han, Y., et al. (2020) Epidemiological and clinical characteristics of 99 cases of 2019 novel coronavirus pneumonia in Wuhan, China: a descriptive study. The Lancet, 395, 507–513.

Diggle, P. J., & Ribeiro, P. J. Jr. (2007) Model-based Geostatistics. Springer, New York (USA).

Dong, Y., Mo, X., Hu, Y., Qi, X., Jian, F., Jiang, F., & Tong, S. (2020) Epidemiology of COVID-19 among children in China. Pediatrics, e20200702, doi: 10.1542/peds.2020-0702.

Ganyani, T., Kremer, C., Chen, D., Torneri, A., Faes, C., Wallinga, J., & Hens, N. (2020) Estimating the generation interval for COVID-19 based on symptom onset data. Preprint, doi:10.1101/2020.03.05.20031815.

Jiang, F., Deng, L., Zhang, L., Cai, Y., Cheung, C. W., & Xia, Z. (2020) Review of the Clinical Characteristics of Coronavirus Disease 2019 (COVID-19). Journal of General Internal Medicine, 35: 15451549.

Lawson, A. B. (2013) Bayesian Disease Mapping: Hierarchical Modeling in Spatial Epidemiology. Second Edition. Boca Rotan: Chapman & Hall.

Neyens, T., Lawson, A. B., Kirby, R. S., and Faes, C. (2016) The bivariate combined model for spatial data analysis. Statistics in Medicine, 35.

R Core Team (2020). R: A language and environment for statistical computing. R Foundation for Statistical Computing, Vienna, Austria. URL https://www.R-project.org/.

Rue, H., Martino, S., & Chopin, N. (2009) Approximate Bayesian inference for latent Gaussian models by using integrated nested Laplace approximations. Journal of the Royal Statistical Society, 71, 319–392.

World Health Organization (WHO)(2020), https://www.who.int/dg/speeches/detail/who-director-general-s-opening-remarks-at-the-media-briefing-on-covid-19-11-march-2020.

Wu F., Zhao S., Yu B., Chen Y.-M., Wang W., Song Z.-G., et al. (2020) A new coronavirus associated with human respiratory disease in China. Nature, 579, 265269.

Yang, W., Cao, Q., Qin, L., Wang, X., Cheng, Z., Pan, A., Dai, J., Sun, Q., Zhao, F., Qu, J., and Yan, F. (2020) Clinical characteristics and imaging manifestations of the 2019 novel coronavirus disease (COVID-19):A multi-center study in Wenzhou city, Zhejiang, China. Journal of Infection, 80, 388–393.

Zhu, N., Zhang, D., Wang, W., Li, X., Yang, B., Song, J. et al. (2020) A novel coronavirus from patients with pneumonia in China, 2019. The New England Journal of Medicine, 382, 727–733. doi: 10.1056/NEJMoa2001017.

